# Age-related clonal hematopoiesis and mosaic sex chromosome loss define distinct systemic proteomic programs and disease vulnerabilities

**DOI:** 10.64898/2026.08.29.26361722

**Authors:** Michael Weyrich, Akshay Ware, Agnes Steixner-Kumar, Johannes Windschmitt, Tamim Sarakpi, Wesley T. Abplanalp, Stefanie Dimmeler, Thimoteus Speer, Andreas M. Zeiher

**Affiliations:** Department of Internal Medicine 4, Goethe-University, Frankfurt, Germany; Else Kroener-Fresenius Center for Nephrological Research, Goethe-University, Frankfurt, Germany; Institute of Cardiovascular Regeneration, Goethe-University, Frankfurt, Germany; Cardio-Pulmonary Institute (CPI), Goethe-University, Frankfurt, Germany; Deutsches Zentrum für Herz-Kreislauf-Forschung (DZHK), partner site Rhine-Main, Frankfurt, Germany

**Author notes:** Corresponding author: Prof. Andreas M. Zeiher, MD, Institute of Cardiovascular Regeneration Goethe-University Frankfurt, Theodor-Stern-Kai 7, 60590 Frankfurt/Main Germany. Both authors contributed equally (Thimoteus Speer, Andreas M. Zeiher).

## Abstract

Clonal hematopoiesis (CH) increases with age, but whether different somatic clones represent an ageing phenotype or exert distinct systemic effects is unclear. In 450,587 UK Biobank participants, including 46,324 with plasma proteomics, we compared clonal hematopoiesis of indeterminate potential (CHIP) and mosaic loss of chromosome Y (mLOY) or X (mLOX) across biological ageing, incident disease, and circulating proteins. Despite shared age dependence, these alterations showed distinct disease spectra: non-DNMT3A CHIP was associated with broad multisystem disease burden, mLOY with a more focused respiratory, musculoskeletal and cardiovascular profile, whereas mLOX lacked broad age-related disease associations. Clone burden mapped to distinct proteomic programs: mLOY to neutrophil degranulation and extracellular-matrix remodeling, non-DNMT3A CHIP to myeloid immune regulation, and mLOX unexpectedly to cytotoxic lymphocyte/NK-cell responses. Mendelian randomization supported selected protein-disease relationships. Thus, age-related hematopoietic clones are not interchangeable markers of ageing but define alteration-specific systemic programs associated with distinct disease vulnerabilities.

## Introduction

Ageing contributes to over 50% of the global disease burden^1^. One of the hallmarks of biological ageing is genomic instability^2^. As humans age, they acquire somatic mutations and genomic alterations affecting all cells in the body^3^. This age-dependent accumulation of somatic genomic alterations results in a somatic mosaicism in multiple cell types and tissues^4^. Most notably, if these acquired genomic alterations occur in hematopoietic stem and progenitor cells and do provide a selective advantage to the cell, in which they occur, this will result in a clonal expansion in the circulating blood. This clonal hematopoiesis (CH) impacts all organs, since the mutated blood cells circulate throughout the body^5^.

The three most common forms of CH associated with ageing are: CH of indeterminate potential (CHIP), defined as the presence of mutations with a variant allele frequency > 2% in genes recurrently altered in hematological diseases in the absence of evidence of malignancy^6^, and sex chromosome aneuploidy, specifically loss of the X chromosome, the most common clonal somatic alteration in leukocytes of females^7^, and loss of the Y chromosome, the most frequent somatic clonal alteration in males^8,9^. These three forms of CH have emerged as risk factors for the development of a variety of age-related diseases and all-cause mortality^10,11^. Specifically, the most frequent CHIP-driver gene mutations in *DNMT3A*, *TET2*, and *ASXL1* are well established to associate not only with cardiovascular diseases, chronic lung diseases, chronic kidney disease, hematologic malignancies and more broadly chronic inflammatory conditions, but also with increased overall mortality^3,12–14^. Likewise, more recent studies also demonstrated that hematopoietic mosaic loss of Y chromosome (mLOY) associates with an increased risk for cardiovascular diseases, general frailty-like ageing phenotypes and chronic fibrosis in lung and kidney as well as decreased longevity^3,15–17^. In contrast, mosaic hematopoietic loss of X chromosome (mLOX) was shown to predominantly affect leukemia risk^18^. While numerous studies addressed the individual associations of these three most common forms of CH with incident diseases, comparative data on how these different forms of CH differentially influence age-dependent clinical outcome are sparse. While some CHIP-driver mutations were previously shown to relate to accelerated epigenetic aging profiles^19,20^, no such data are currently available for mLOX and mLOY in large population-based cohorts. More importantly, information on the plasma protein profile associated with the presence of these subclinical clonal somatic alterations is very limited. In unbiased, population-wide screens, CHIP and mLOY have been previously reported to associate with specific proteomic signatures implicated in cardiovascular diseases^21,22^, while the effects of mLOX on circulating plasma proteins have only been studied in very limited and selected sample sizes of circulating biomarkers^23^.

Thus, using the population-wide data of the UK biobank, it was the aim of the present study to address the differential effects of CHIP, mLOX and mLOY on 1) their individual association with accelerated aging as measured by biomarker-derived age estimates and proteomic aging clocks, 2) their differential contribution to incident common non-malignant, age-related diseases, 3) their individual additive information to a general proteomic ageing clock to predict the risk of common age-related diseases, and finally 4) to identify CH-specific plasma proteomic profiles and their relation to common age-associated diseases with support for genetic causal inference from Mendelian randomization in an external cohort.

## Results

### Association of mLOX, mLOY, and CHIP with age

We first quantified mLOX, mLOY, and CHIP in UK Biobank participants with available genomic data. Among 502,501 UK Biobank participants, 450,587 had quantification of mLOX, mLOY, and CHIP. 426,882 had incident disease outcomes and complete covariate information, and 46,324 had OLINK plasma proteomic profiling available for downstream proteomic analyses (**Fig. 1A**, **Supplementary Table 1**). In men, mLOY (cut-off ≥10%) was more prevalent than CHIP (cut-off VAF ≥2%), whereas mLOX (cut-off ≥5%) in women was substantially less frequent than mLOY in men (**Fig. 1B**). Co-occurrence of mLOY with CHIP was only detectable in a small number of participants. All three forms of CH exponentially increased with age, with mLOY showing the steepest age-related increase (**Fig. 1C**).

**Figure 1.**
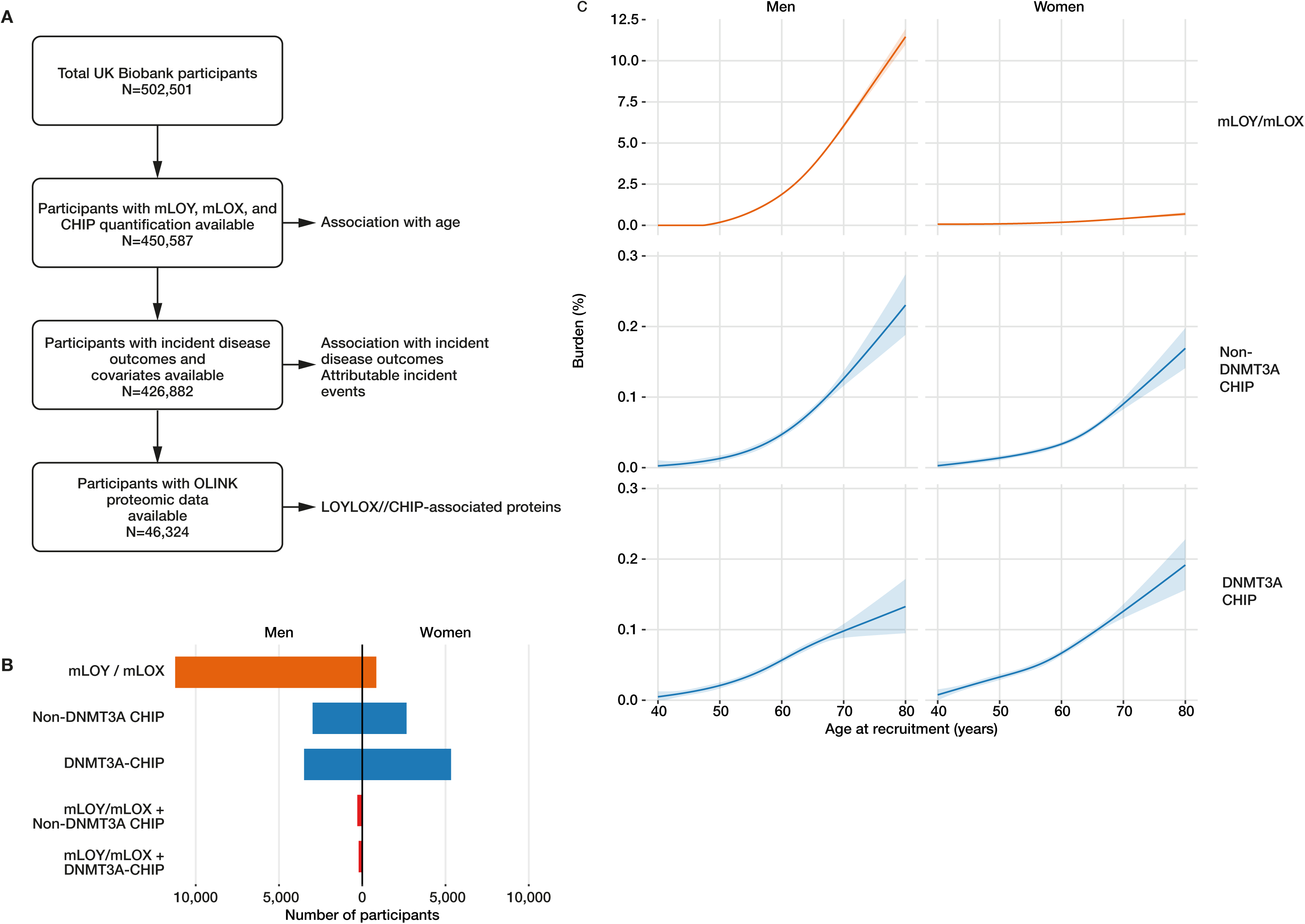
Study population, prevalence and age-related burden of mosaic sex-chromosome loss and CHIP. **(A)** Flow diagram of the UK Biobank analysis population. Of 502,501 participants, 450,587 had mLOY, mLOX or CHIP quantification, 426,882 had incident-disease outcomes and complete covariate data, and 46,324 had Olink plasma-proteomic data. **(B)** Numbers of men and women with mLOY or mLOX, non-DNMT3A CHIP, DNMT3A-CHIP, or co-occurring sex-chromosome loss and CHIP. Clinical thresholds were mLOY ≥10%, mLOX ≥5% and CHIP VAF ≥2%. **(C)** Adjusted age-dependent clone burden from ages 40–80 years, shown separately in men and women. Curves were estimated with natural cubic splines of age (four degrees of freedom) and adjusted for smoking, ethnicity, BMI, LDL cholesterol, hypertension, diabetes, and genetic principal components 1-10. mLOY and mLOX are shown as estimated leukocyte percentages, and non-DNMT3A and DNMT3A-CHIP as maximum VAF. Shading denotes 95% confidence intervals. LOY, loss of chromosome Y; LOX, loss of chromosome X; VAF, variant allele fraction.

### mLOY and CHIP increase the risk for the first occurrence of common age-related diseases, associate with all-cause mortality and accelerated biological ageing estimates

Since age is the central driver of disease susceptibility, we next examined whether mLOX, mLOY, and CHIP were associated with an increased risk for the first occurrence of common age-related diseases and all-cause mortality. CHIP was a priori modeled as a non-DNMT3A- CHIP composite comprising all CHIP-driver mutations excluding DNMT3A, whereas DNMT3A-CHIP-driver mutations were treated as a separate pathogenic driver. Common age-related diseases were defined as ischemic heart disease, heart failure, stroke, Alzheimer’s disease, pneumonia, chronic obstructive pulmonary disease (COPD), sepsis, chronic kidney disease, type-2 diabetes, osteoporosis, and polyarthritis. In fully adjusted Cox models, non-DNMT3A-CHIP in both, women and men, and mLOY in men significantly increased the hazard ratios for the risk for the first occurrence of any of these age-related diseases as well as for all-cause mortality. In men, DNMT3A-mutant CHIP was not associated with the first occurrence of age-related diseases, but was associated with all-cause mortality. In women, weak but statistically significant associations were observed for both the first occurrence of age-related diseases and all-cause mortality **(Fig. 2A, Supplementary Table 2)**. Interestingly, mLOX did not show robust associations with either an increased risk for the first occurrence of age-related diseases nor with all-cause mortality in the fully adjusted models (**Fig. 2A**, **Supplementary Table 2**).

**Figure 2.**
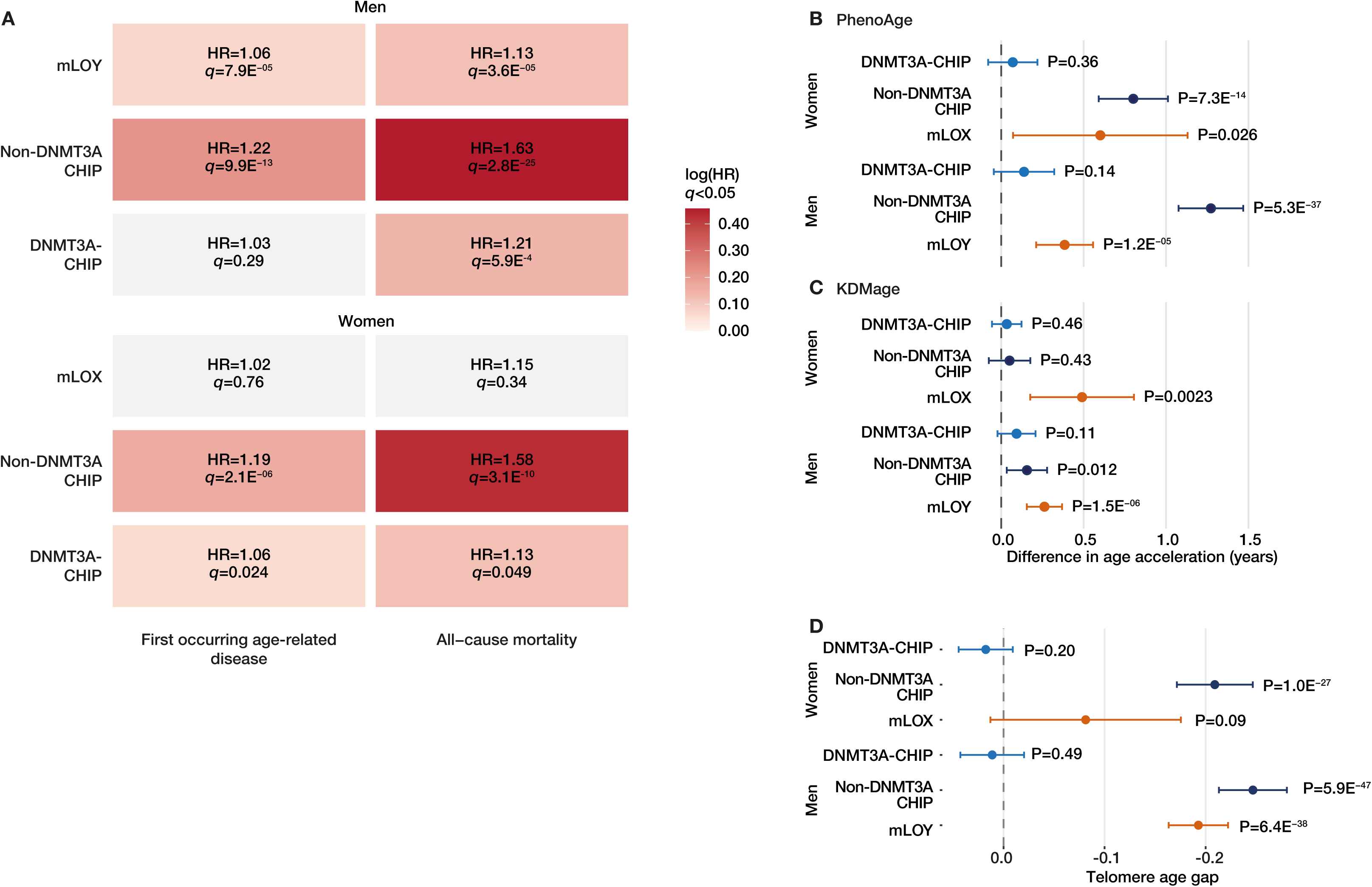
Associations with age-related disease, mortality, and biological-age measures. **(A)** Sex-stratified hazard ratios for the first recorded post-baseline incident event among 11 age-related disease groups and for all-cause mortality. Fully adjusted Cox models included age at recruitment, smoking, ethnicity, BMI, LDL cholesterol, hypertension, prevalent diabetes, and genetic principal components 1–10. Follow-up was truncated at 10 years. *q* values were calculated by the Benjamini-Hochberg method within each outcome and model across the six sex-exposure tests. (**B–D**) Adjusted differences in PhenoAge acceleration **(B)**, Klemera–Doubal method age acceleration **(C)** and telomere age gap **(D)**. Points and bars denote regression coefficients and 95% confidence intervals. HR, hazard ratio; KDM, Klemera-Doubal method.

Landmark analyses across 0-3, 3-6 and 6-10 years supported the temporal robustness of the principal clinical associations. Non-DNMT3A CHIP remained associated with all-cause mortality in men and women in each interval. Associations with the first occurrence of an age-related disease were also consistently elevated in women and increased numerically across intervals in men. For mLOY, associations were close to the null during the first three years, but were evident for both outcomes after year 3. None of the formal time-window heterogeneity tests remained significant after outcome-specific false-discovery-rate correction (**Supplementary Table 3**).

To place these findings in the context of biological ageing, we investigated how mLOX, mLOY, and CHIP interfere with established biomarker-derived biological age estimates and telomere-age gap. mLOX, mLOY, and non-DNMT3A-CHIP were significantly associated with increased PhenoAge acceleration (**Fig. 2B, Supplementary Table 4**). Likewise, mLOY and non-DNMT3A-CHIP in men as well as mLOX showed significant associations with KDMage acceleration (**Fig. 2C, Supplementary Table 4**). There were no significant associations between DNMT3A-CHIP and these measures of biological ageing (**Fig. 2B-C**). Lastly, mLOY in men and non-DNMT3A-CHIP in both sexes were significantly associated with a lower telomere length age gap indicating shorter telomeres than expected for age (**Fig. 2D, Supplementary Table 4**). Together, these results indicate that age-related clonal hematopoietic alterations differ substantially in their associations with biological ageing and clinical outcomes. While mLOY and non-DNMT3A CHIP were associated with both accelerated biological ageing and increased age-related disease risk, mLOX was associated with biological age acceleration without a corresponding increase in disease incidence or mortality.

### mLOY and CHIP show distinct disease-burden signatures in men

We next characterized the disease-wide association profiles of mLOY, non-DNMT3A CHIP and DNMT3A-mutant CHIP in men. Across ICD-coded outcomes, CHIP showed broad associations spanning hematological, circulatory, infectious, genitourinary, and respiratory disease chapters, whereas mLOY showed a more focused pattern involving respiratory, infectious, musculoskeletal, and circulatory disease domains (**Fig. 3A-B and Supplementary Table 5**).

**Figure 3.**
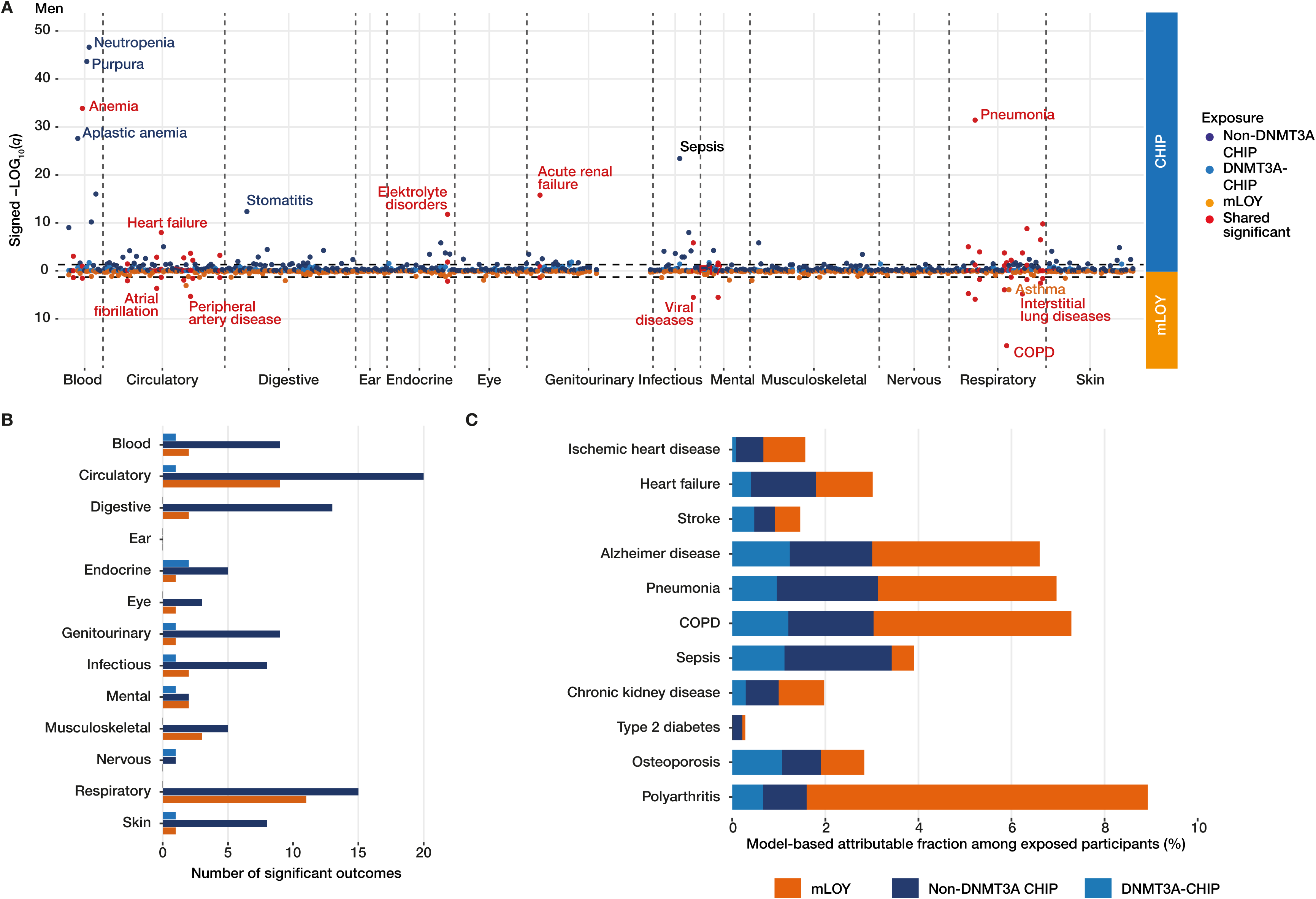
Phenome-wide disease associations and exposure-specific model-based attributable fractions in men. **(A)** Miami plot of incident ICD-10 outcomes associated with mLOY, non-DNMT3A CHIP or DNMT3A-CHIP. The vertical axis shows signed −LOG_10_(*q*): CHIP results are displayed above zero and mLOY results below zero. Colors distinguish the three exposures and outcomes significant for both mLOY and either CHIP exposure. Selected outcomes are labelled. Benjamini-Hochberg *q* values were calculated across all eligible outcomes within each exposure, and *q*<0.05 defined significance. Only outcomes with at least 200 incident events were tested. **(B)** Number of significant outcomes by ICD-10 chapter. **(C)** Exposure-specific model-based attributable fractions among exposed participants (AF) for 11 selected age-related diseases. AF represents the proportion of predicted 10-year events among participants carrying the indicated exposure that was associated with that exposure under the respective fully adjusted Cox model.

For the 11 selected age-related diseases, model-based attributable fractions among exposed participants (AF) differed across exposures and outcomes. Among men with mLOY, the largest AF estimates were observed for polyarthritis, COPD, pneumonia and Alzheimer’s disease, whereas among men with non-DNMT3A CHIP the largest estimates were observed for sepsis, pneumonia, COPD and Alzheimer’s disease. DNMT3A-mutant CHIP generally showed smaller AF estimates (**Fig. 3C and Supplementary Table 7**). Together with the phenome-wide association patterns, these findings indicate that mLOY, non-DNMT3A CHIP and DNMT3A-mutant CHIP are associated with partially distinct disease profiles.

### CHIP dominates the attributable disease burden in women

In women, the phenome-wide analysis revealed broad associations of CHIP across ICD chapters, whereas mLOX was not robustly associated with incident disease outcomes after correction for multiple testing (**Fig. 4A-B and Supplementary Table 6**). Non-DNMT3A CHIP showed associations across hematological, circulatory, digestive, infectious, and respiratory disease categories.

**Figure 4.**
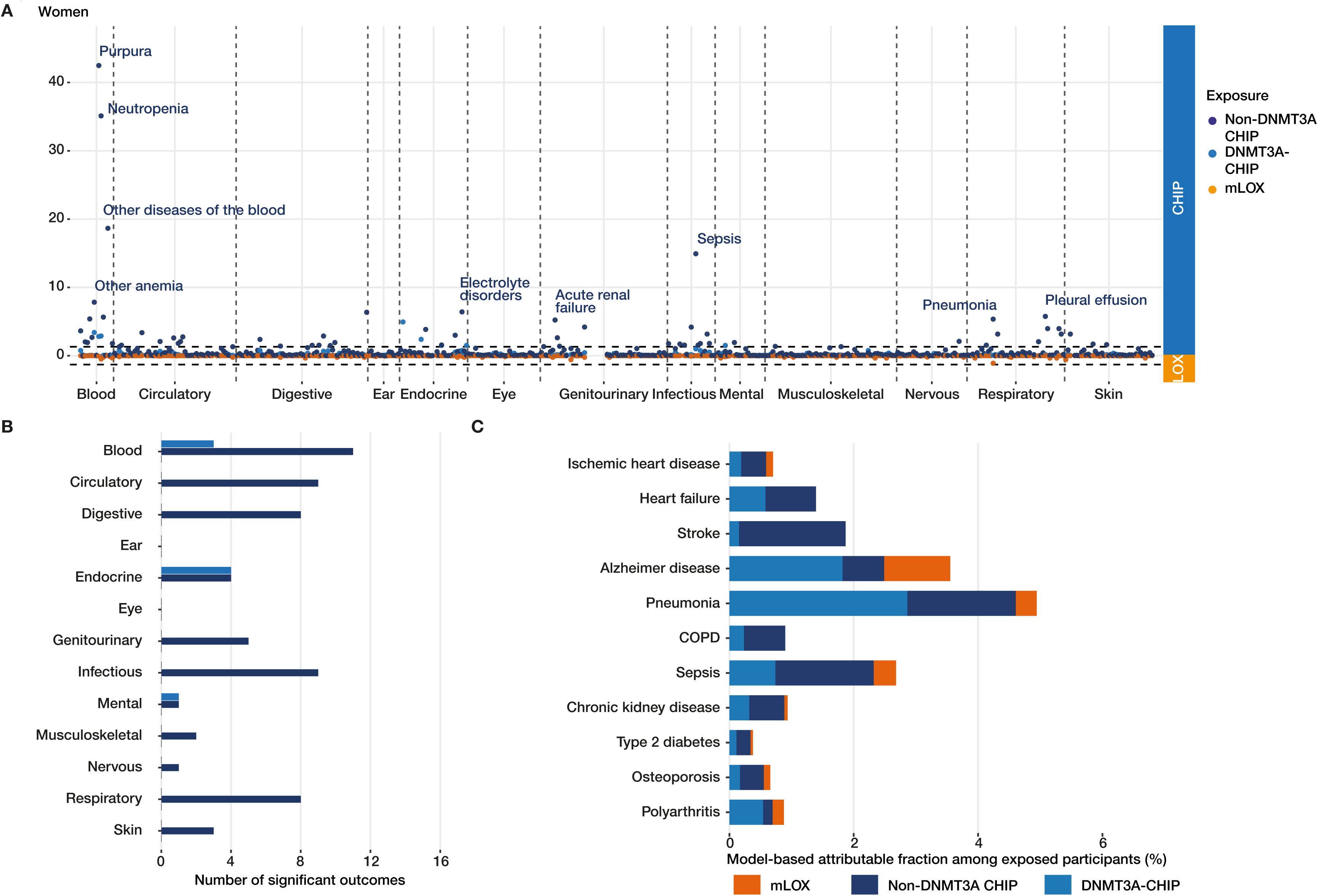
Phenome-wide disease associations and exposure-specific model-based attributable fractions in women. **(A)** Miami plot of incident ICD-10 outcomes associated with mLOX, non-DNMT3A CHIP or DNMT3A-CHIP. The vertical axis shows signed −LOG_10_(*q*): CHIP results are displayed above zero and mLOX results below zero. Colors distinguish the exposures. Selected outcomes are labelled. Benjamini-Hochberg *q* values were calculated across all eligible outcomes within each exposure, and *q*<0.05 defined significance. Only outcomes with at least 200 incident events were tested. **(B)** Number of significant outcomes by ICD-10 chapter. **(C)** Exposure-specific model-based attributable fractions among exposed participants (AF) for 11 selected age-related diseases. AF represents the proportion of predicted 10-year events among participants carrying the indicated exposure that was associated with that exposure under the respective fully adjusted Cox model.

Among women carrying non-DNMT3A CHIP, the largest model-based AF estimates were observed for pneumonia, stroke, sepsis, heart failure and Alzheimer’s disease. Among women carrying DNMT3A-mutant CHIP, the largest estimates were observed for pneumonia and Alzheimer’s disease. AF estimates for mLOX were generally smaller and were observed for only selected endpoints (**Fig. 4C and Supplementary Table 7**). These findings indicate that CHIP has a broader disease-association profile than mLOX in women.

### mLOY and CHIP add disease risk beyond proteomic ageing

Because mLOY, mLOX and non-DNMT3A-CHIP were associated with accelerated ageing phenotypes, we next investigated whether these exposures added risk to predict incident age-related diseases beyond a recently published and comprehensively validated proteomic ageing score^24^. Participants with an elevated proteomic ageing score (above median) had an increased cumulative incidence of age-related diseases. In men, the addition of mLOY or non-DNMT3A-CHIP to a high proteomic ageing score further significantly separated cumulative incidence curves for age-related diseases, indicating that mLOY and non- DNMT3A-CHIP capture disease-relevant biology beyond accelerated proteomic ageing alone (**Fig. 5A**, **Supplementary Table 8**). In fact, the addition of mLOY or non-DNMT3A-CHIP approximately doubled the 10-year cumulative incidence of age-related diseases compared to the proteomic age clock alone over a 10-year follow-up period. In women, non-DNMT3A- CHIP also showed a slight additive effect on top of an elevated protein age clock, whereas mLOX did not further increase the cumulative occurrence of incident age-related diseases (**Fig. 5A**).

**Figure 5.**
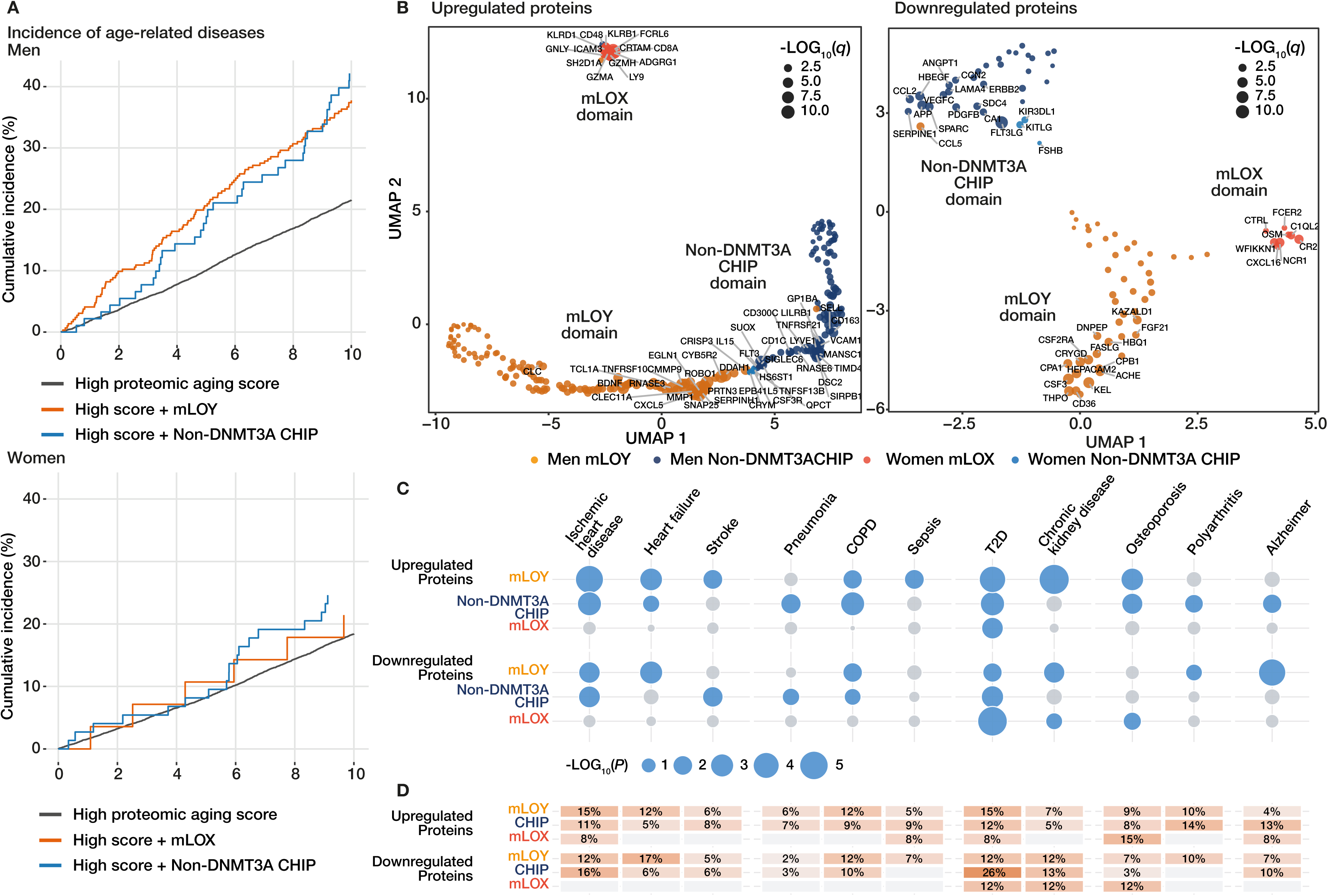
Proteomic ageing, protein-domain landscapes, and genetic disease evidence. **(A)** Cumulative incidence of the 11-disease age-related composite among men and women with a high 20-protein age gap, defined as at or above the sex-specific median. Curves show high proteomic age gap alone, with mLOY or mLOX, or with non-DNMT3A CHIP. Participants with any prevalent component disease were excluded. Cumulative incidence was estimated as one minus the Kaplan-Meier survival estimate over 10 years. **(B)** UMAP representations of proteins upregulated or downregulated in association with continuous mLOY, mLOX or non-DNMT3A CHIP burden. Each point is a protein. Color denotes the dominant sex-exposure profile and size denotes −LOG_10_(*q*). **(C)** Outcome-specific aggregated Cauchy association test (ACAT) results combining primary protein-to-disease Mendelian-randomization *P* values within each upregulated or downregulated domain in FinnGen. Bubble area is proportional to −LOG_10_(*P*). Blue denotes nominal *P*<0.05 and grey *P*≥0.05. **(D)** Percentage of proteins in each domain with a nominal primary inverse-variance-weighted or Wald-ratio *P*<0.05 for each FinnGen outcome. COPD, chronic obstructive pulmonary disease; CKD, chronic kidney disease; T2D, type 2 diabetes; IVW, inverse-variance weighted.

These data indicate that mLOX, mLOY and non-DNMT3A-CHIP do not only reflect an unspecific ageing-associated protein profile, but point towards specific proteins altered by mLOX, mLOY and non-DNMT3A-CHIP. Therefore, using the extent of mLOX, mLOY and the VAF of CHIP-driver mutations as continuous variables in order to avoid potential bias by the selection of cut-off values, we identified circulating plasma proteins being significantly up- or down-regulated in association with the extent of the different CH (**Supplementary Table 9**). We then mapped the CH-associated proteins onto a UMAP representation of the plasma proteome, separating upregulated and downregulated proteins (**Supplementary Table 11**).

As illustrated in **Figure 5B**, proteins associated with mLOX, mLOY and non-DNMT3A-CHIP occupied in large part distinct domains for both, up- and down-regulated proteins. DNMT3A- CHIP did only show very few statistically significantly regulated proteins. These findings indicate that mLOX, mLOY, and non-DNMT3A-CHIP are associated with distinct proteomic signatures rather than a single shared ageing-related protein pattern (**Fig. 5B**).

In sensitivity analyses additionally adjusted for circulating neutrophil, lymphocyte, monocyte, eosinophil and basophil percentages and platelet count, 194 mLOY-, 19 mLOX-, 156 male non-DNMT3A CHIP- and 12 female non-DNMT3A CHIP-associated proteins remained significant at a false-discovery rate below 0.05. Compared with models fitted in the same blood-count-complete participants without these additional covariates, protein-effect profiles remained strongly correlated (Pearson r=0.903, 0.907, 0.981 and 0.989, respectively). (**Supplementary Table 10**).

### Proteomic domains link mLOY, mLOX and CHIP to age-related disease

To assess whether the CH-associated proteomic domains were genetically linked to age-related diseases, we used protein-to-disease Mendelian randomization with disease outcomes from the independent FinnGen cohort. For each CH-specific proteomic domain, we then combined the Mendelian-randomization *P* values across its constituent proteins using the aggregated Cauchy association test. At a nominal threshold of *P*<0.05, the mLOY- upregulated and non-DNMT3A CHIP-upregulated domains each showed aggregate genetic evidence for 8 of the 11 age-related disease outcomes, although their outcome profiles were only partly overlapping (**Fig. 5C**, **Supplementary Table 12**). The mLOY-downregulated and non-DNMT3A CHIP-downregulated domains showed corresponding signals for seven and five outcomes, respectively. By contrast, the mLOX-upregulated domain showed evidence only for type 2 diabetes, whereas the mLOX-downregulated domain showed evidence for type 2 diabetes, chronic kidney disease and osteoporosis. Thus, genetic support for protein-disease associations was broader across the mLOY- and non-DNMT3A CHIP-related domains than across the mLOX-related domains. Because ACAT aggregates evidence across proteins without estimating a common domain-level effect size or direction, these findings prioritize domain-disease relationships, but do not establish a causal effect of an entire proteomic domain. At the individual-protein level, the proportion of proteins showing nominal primary Mendelian-randomization associations varied across domains and disease outcomes, providing complementary protein-level support for selected domain-disease relationships (**Fig. 5D**, **Supplementary Table 13-14**).

### Functional annotation identifies immune, extracellular-matrix and hematopoietic signaling programs

Finally, we functionally annotated the proteins defining the mLOX-, mLOY- and non- DNMT3A-CHIP-associated proteomic domains. Objective annotation identified six association-profile clusters: hematopoietic growth-factor signaling, neutrophil degranulation and extracellular matrix remodeling, myeloid immune regulation, chemotactic, hemostatic and extracellular-matrix signaling, cytotoxic lymphocyte/NK-cell response and B-cell/cytokine-associated signaling (**Fig. 6A** and **Supplementary Tables 15 and 16**). Support was strongest for the myeloid immune-regulatory, chemotactic/hemostatic/extracellular-matrix and cytotoxic lymphocyte/NK-cell annotations, moderate for the neutrophil-degranulation annotation, and less robust for the hematopoietic growth-factor and B-cell/cytokine-associated annotations. The latter was treated as exploratory. In men, mLOY and non-DNMT3A CHIP showed opposing, cluster-specific protein regulation. mLOY was characterized by upregulation of the neutrophil-degranulation and extracellular matrix remodeling profile and downregulation of hematopoietic growth-factor signaling (**Fig. 6B**). Non-DNMT3A CHIP was preferentially associated with upregulated myeloid immune regulation and B-cell/cytokine-associated signaling and downregulated chemotactic, hemostatic and extracellular-matrix signaling. TET2- and ASXL1-mutant CHIP, which accounted for 2/3 of the non-DNMT3A-CHIP participants, showed comparable patterns (**Supplementary Figure 1A**), with ASXL1 additionally associated with upregulation of selected vascular-health-promoting enzymes, as previously reported^21^. In women, non- DNMT3A CHIP showed weaker myeloid and cytokine-associated immune signals. Most notably, mLOX was distinguished by a very peculiar and characteristic protein signature with upregulation of a cytotoxic lymphocyte/NK-cell response and downregulation of B-cell/cytokine-associated signaling (**Fig. 6C**).

**Figure 6.**
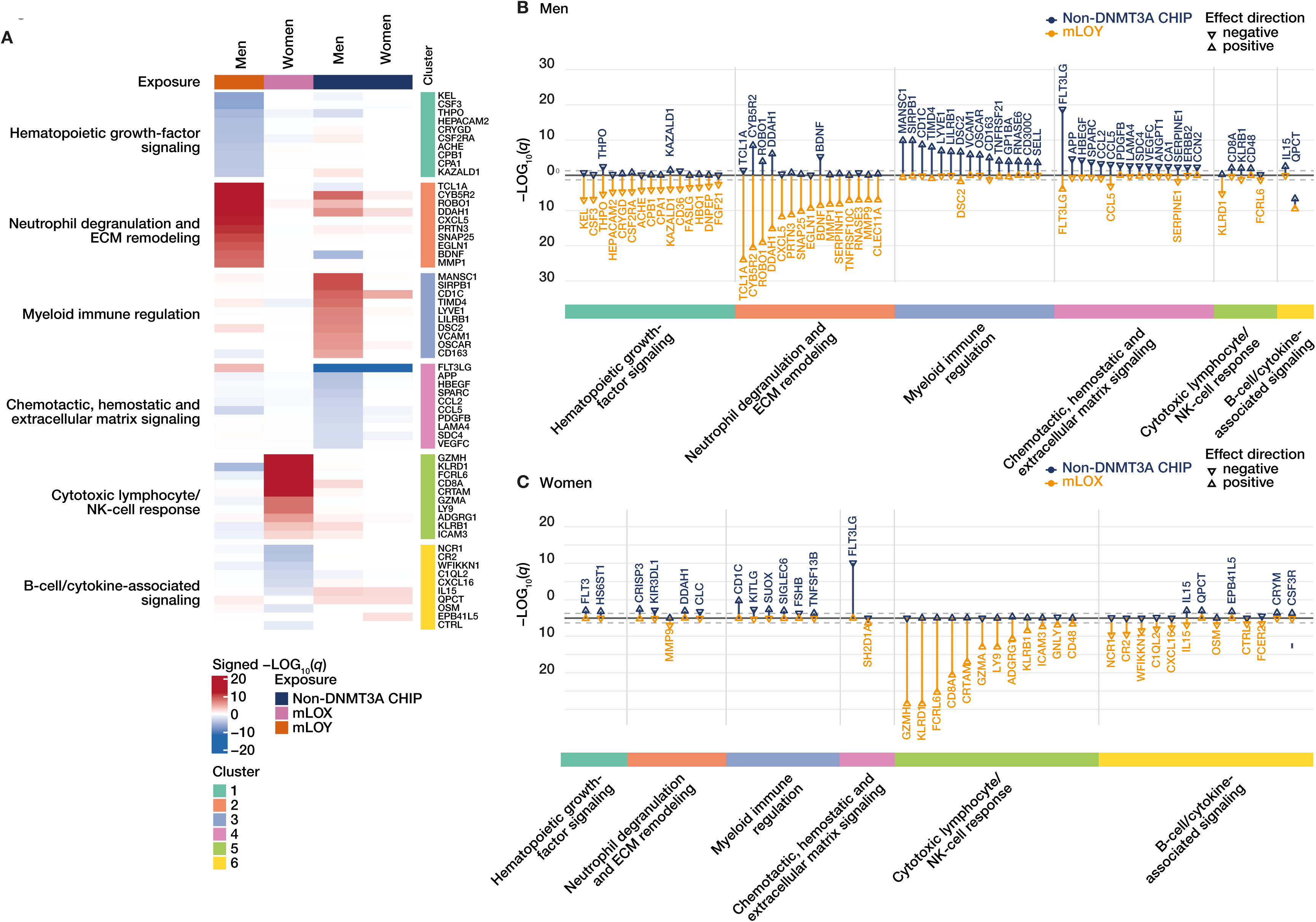
Distinct functional protein programs associated with mLOY, mLOX and non-DNMT3A CHIP. **(A)** Heatmap of representative proteins from six association-profile clusters: haematopoietic growth-factor signalling, neutrophil degranulation and extracellular-matrix remodelling, myeloid immune regulation, chemotactic, hemostatic and extracellular-matrix signalling, cytotoxic lymphocyte/NK-cell response and B-cell/cytokine-associated signalling. Columns show mLOY, mLOX and non-DNMT3A CHIP. Values are signed −LOG10(*q*), with red indicating positive and blue negative associations. Up to ten proteins per cluster with *q*<0.05 in at least one profile are shown. Cluster labels were assigned using the enrichment and stability framework described in Methods. **(B)-(C)** Sex-specific mirror plots for men (**B**; non-DNMT3A CHIP above zero and mLOY below zero) and women (**C**; non-DNMT3A CHIP above zero and mLOX below zero). Vertical position separates the two exposure profiles for display and its magnitude is −LOG10(*q*). Triangle orientation indicates the sign of the regression coefficient. Dashed lines mark *q*=0.05. ECM, extracellular matrix.

## Discussion

The results of the present study significantly extend previous reports addressing the role of clonal somatic hematopoietic alterations for incident age-associated diseases and disclose specific differential circulating proteomic profiles encoding clinically relevant disease-related information.

In line with results of prior studies, the occurrence of all investigated CH significantly increases with age, with mLOY and non-DNMT3A-CHIP showing the steepest age-related increase in men, whereas CHIP-driver mutations dominate CH in women compared to mLOX. However, the three forms of CH differentially contribute to both, the incidence of prototypical age-associated diseases as well as to the overall disease burden. While non- DNMT3A-CHIP significantly associated with age-associated diseases as well as all-cause mortality in both women and men, only mLOY in men, but not mLOX in women correlated with incident age-associated diseases and all-cause mortality. DNMT3A-CHIP was only weakly associated with incident diseases or mortality in both sexes. Importantly, the differentially increased risk for incident age-related diseases was parallelled by differences in biological age acceleration as measured by the biomarker-derived biological age estimates PhenoAge or KDMage as well as the cellular marker of an increased telomere gap. These data extend previous reports showing associations of CHIP with accelerated epigenetic aging as would be expected, since DNMT3A, TET2 and ASXL1 represent epigenetic modifiers^19^. The present data show that non-DNMT3A-CHIP, mLOX and mLOY are significantly related to accelerated biological aging as measured by biomarker-derived age estimates, while DNMT3A-CHIP did not associate with accelerated biological aging or increased telomere gap.

In men, non-DNMT3A-CHIP and mLOY contributed to a large number of significant disease outcomes, yet mLOY demonstrated a more focused pattern with preferential associations with circulatory, respiratory, and musculoskeletal disease entities. In contrast, in women, the broad disease-wide burden revealed a predominant contribution of non- DNMT3A-CHIP exposure, with no significant contribution of mLOX. Notably, DNMT3A-CHIP was associated with very few incident disease patterns including some forms of infection, osteoporosis, and polyarthritis in both sexes, confirming previous reports^25–28^. These data indicate that the three forms of age-related CH are not interchangeable age-associated markers, but rather confer partially distinct disease liabilities. Moreover, as previously reported, non-DNMT3A-CHIP-driver mutations associate with different disease burdens compared to DNMT3A-CHIP-driver mutations^29^.

Circulating proteins may offer a promising avenue not only for assessing biological age, but also for deciphering their potential specific contribution to different disease susceptibilities due to their direct involvement in biological functions^30^. It is well established that proteomic age associates with multimorbidity^24,31^ and – more importantly – predicts the first occurrence of incident diseases^32,33^. Therefore, it has been proposed to use protein aging clocks to quantify a general vulnerability for accumulating diseases^33^. The results of the present study now disclose that non-DNMT3A-CHIP and mLOY in men as well as – to a lesser extent – non-DNMT3A-CHIP, but not mLOX in women significantly and profoundly increased the risk to predict the incidence of age-related diseases, when added to a recently published and extensively validated 20-protein aging score^24^. These data demonstrate that non-DNMT3A-CHIP, mLOX and mLOY do not only reflect an unspecific age-associated protein profile, but their individual circulating protein signatures point towards specifically altered proteins with involvement in biological functions to contribute to the differential susceptibility to age-related diseases. Indeed, both, up- and down-regulated proteins significantly correlating with the extent of individual CH alterations in blood clustered to individual CH domains. These findings support our hypothesis that the circulating proteomic signatures of mosaic hematopoietic sex chromosome aneuploidy and CHIP are not only age-related exposure profiles, but may also encode clinically relevant information for disease susceptibility. Most notably, support for genetic causal inference from Mendelian randomization in a large external cohort, the FinnGen database, revealed that CH variables associate with changes in a number of proteins, which are causally related to a variety of the age-related diseases investigated.

Functional annotation of the individual proteins defining the specific CH domains provided important insights into the mechanisms involved in the vulnerability for specific age-related diseases. Among the top regulated proteins (by significance), mLOY was associated with selective upregulation of proteins involved in neutrophil degranulation and extracellular matrix remodeling, including the chemokine CXCL5 and the neutrophil serine protease PRTN3, both experimentally and clinically implicated to contribute to lung fibrosis and COPD^34,35^, as well as the matrix remodeling proteins ROBO1^36^, MMP1^37^, SERPINH1, which encodes HSP47^38,39^, and MMP9^40^, which all have been shown to associate with diffuse organ fibrosis. On the other hand, mLOY associated with significant downregulation of proteins involved in hematopoietic growth-factor signaling, including CD36, a master regulator of immune cell metabolism and proinflammatory cytokine release^41,42^, FASLG, which inhibits cytotoxic T cells^43^, and FGF21, essential for metabolic homeostasis and experimentally shown to increase life span in mice^44,45^. These findings offer important proteomic insights into potential mechanisms involved in the well-established and prominent role of mLOY to preferentially associate with diffuse chronic fibrotic diseases like heart failure^16,46^, chronic kidney disease^47,48^, COPD^17,49,50^ and other diffuse fibrotic, age-related organ diseases^3^. In addition, the present study further confirms our previous report, that the top mLOY- upregulated protein was TCL1A, an Akt-co-activator implicated to play a role for the genetic susceptibility to mLOY, which may also confer a proliferation benefit and, thus, clonal expansion for LOY hematopoietic cells^9,22^.

In contrast, non-DNMT3A-CHIP in men was most prominently associated with profound upregulation of proteins involved in myeloid immune regulation, including CD1C and TIMD4, which both activate immune cells^51,52^, the direct activating receptors on myeloid cells SIRPB1^53^ and CD300c^54^, and LILRB1, VCAM1, OSCAR, CD163 and SELL, the circulating protein levels of which are indicative of an increased systemic inflammatory activity^55–59^. These findings indicate a persistent low-grade inflammatory activation mediated in large part by activated immune cells to associate with non-DNMT3A-CHIP, which is in line with numerous experimental studies modeling the effects of TET2- or ASXL1-CHIP-driver mutations in hematopoietic stem cells^3,29^. In the present proteomic study, >65% of the non- DNMT3A-CHIP participants were carriers of either TET2- or ASXL1-CHIP-driver mutations. Notably, in multiple ways, these findings considerably extend the results of the only previously published report on the proteomic profile of non-DNMT3A-CHIP showing an enrichment in inflammation and immune response pathways^21^. First, for assessing the association between circulating proteins and CHIP-driver mutations, we did use the extent of driver mutations (variant allele frequencies) as a continuous variable rather than a fixed cut- off in order to avoid potential bias by the selection of an arbitrary threshold (e.g. >2% for CHIP definition or >10% for large clones). Second, we analyzed the association of the CHIP-related protein signatures to a large range of age-related diseases rather than focusing only on cardiovascular diseases. Third, we evaluated the disease relevance of the non-DNMT3A CHIP-associated proteomic domains using external two-sample Mendelian randomization based on protein-associated genetic instruments and FinnGen disease-outcome summary statistics. This analysis tested whether genetically predicted protein levels were associated with disease risk in the large FinnGen cohort. Taken together, the protein signature reflecting low-grade chronic inflammation associated with non-DNMT3A-CHIP underscores the role of CHIP as a risk factor for the incidence of a broad range of low-grade chronic inflammatory diseases, to which the prototypical age-related diseases all belong^60^.

The most striking observation of the present study relates to the hitherto unreported specific association of mLOX with upregulated proteins involved in cytotoxic lymphocyte/NK-cell responses. The cell-surface immune cell receptors KLRD1, also known as CD94, a NK cell sensor to regulate cytotoxic immune responses, FCRL6, expressed on NK cells and cytotoxic T cells, LY9, also known as CD229, as well as ADGRG1, also known as GPR56 and expressed on cytotoxic lymphocytes, have all been implicated in different leukemic diseases^61–64^. Moreover, the serine protease effector molecules granzyme A and H released from cytotoxic immune cells to mediate cytotoxic killing as well as CRTAM and CD8A as well-established markers of activated cytotoxic T cell lineages are all known to contribute to autoimmunity and tissue destruction^65–68^. Taken together, these findings define a specific circulating protein signature, which mechanistically supports not only previous studies showing a prominent role for mLOX to clinically associate with myeloid and lymphoid leukemias, but also genetic analyses identifying genetic variants in numerous genes associated with autoimmune diseases to influence the development of mLOX^18^. Importantly, this cytotoxic lymphocyte/NK-cell signature is consistent with genetic studies of mLOX. Large-scale genetic analyses identified mLOX-associated variants at multiple immune-related loci, with enrichment of associations related to immunity and HLA biology^18^. These findings suggest that immune-mediated selection may contribute to the emergence or expansion of X-loss clones and provide independent genetic support for our observation that mLOX is associated with a distinct cytotoxic lymphocyte/NK-cell response. Thus, while mLOX is an age-related phenomenon and associates with accelerated biological aging as measured by biomarker-derived biological age estimates, its circulating protein signature does not relate to prototypical age-associated diseases providing a rational explanation for the lack of mLOX to correlate with an increased risk for incident age-related diseases.

Several limitations should be considered. First, this was an observational analysis of a volunteer cohort, and the UK Biobank population is healthier and less ethnically diverse than the general population. Although the association models adjusted for major demographic, clinical and genetic covariates, residual and unmeasured confounding cannot be excluded, and the reported associations and model-based attributable fractions should not be interpreted as causal effects. Second, mLOY, mLOX and CHIP were measured at a single time point. The analyses therefore did not capture subsequent clone emergence, expansion or regression. However, our landmark analyses supported the temporal robustness and directionality of the principal clinical associations over time and none of the formal time-window heterogeneity tests remained significant after outcome-specific false-discovery-rate correction Third, plasma proteins were measured cross-sectionally in a smaller subset of the cohort. Consequently, temporal ordering between clonal alterations and protein changes cannot be established. Moreover, because clonal alterations in hematopoietic stem and progenitor cells are expected to modify the abundance of circulating leukocyte lineages, the primary proteomic analyses were a priori not adjusted for cell counts in order to preserve potential clone-driven compositional biology as part of the exposure signal. However, sensitivity analyses additionally adjusting for blood cell composition revealed that the overall protein-effect profiles were highly concordant with the primary model and the six-domain proteomic architecture was preserved with >95% of the proteins retaining their corresponding biological domain assignment after adjustment for circulating blood-cell composition. Finally, Mendelian randomization provides orthogonal genetic support, but relies on instrument relevance, independence and exclusion-restriction assumptions.

In summary, the specific circulating protein signatures associated with the three most common different hematopoietic CH alterations do not only provide important insights into the mechanisms involved in the differential susceptibility to incident age-related diseases, but the prioritized pathways may also offer specific precision-guided therapeutic avenues to interfere with the biological functional sequelae of harboring age-related somatic mutations or sex chromosomal aneuploidy in circulating blood cells.

## Methods

### Study design and participants

This study used the prospective UK Biobank cohort, which has been described previously^69^. UK Biobank recruited 503,317 volunteers aged 40–69 years at 22 assessment centers in England, Scotland and Wales between 2006 and 2010. Baseline questionnaire, physical-measurement and biological-sample data were linked to longitudinal health records and mortality data. The analysis data set contained 502,501 participants. 450,587 had data permitting ascertainment of mosaic loss of chromosome Y (mLOY), mosaic loss of chromosome X (mLOX) or clonal haematopoiesis of indeterminate potential (CHIP). 426,882 had incident-outcome and complete covariate data for the principal clinical analyses, and 46,324 had plasma proteomic data. Analysis-specific sample sizes therefore varied according to sex, exposure availability, outcome availability and complete covariate information, as summarized in Fig. **1A**.

Analyses were stratified by UK Biobank-recorded sex because mLOY and mLOX are sex-chromosome-specific exposures. Participants coded as male were included in mLOY analyses and those coded as female in mLOX analyses. All eligible participants with the data required for each analysis were included. Except where explicitly stated for proteomic measurements, missing data were handled by complete-case analysis.

### Ethics

UK Biobank operates with approval from the North West Multi-center Research Ethics Committee (11/NW/0382), and all participants provided informed consent. The present analyses were conducted under UK Biobank application 170256.

### Ascertainment of mosaic loss of chromosome Y and X

Mosaic sex-chromosome loss in peripheral-blood DNA was called from genotyping-intensity data using the Mosaic Chromosomal Alterations (MoChA) pipeline^70^. The same MoChA pipeline was used to call mLOY in men and mLOX in women. Clone extent was expressed as the estimated percentage of affected leukocytes. For the analyses, mLOY was defined as a clone extent of at least 10% in men and mLOX as at least 5% in women^18,71^. For continuous proteome-wide analyses, clone extent was transformed as LOG(1+percentage) and standardized within sex.

### Ascertainment of clonal hematopoiesis of indeterminate potential

CHIP status and variant allele fractions (VAFs) were obtained from a curated UK Biobank whole-exome sequencing call set. The source workflow identified candidate somatic variants in recurrently mutated hematological driver genes and applied sequencing-quality, functional-annotation and population-based filters to reduce technical artefacts and residual germline variation. CHIP was defined at VAF ≥2%. DNMT3A-mutant CHIP was analyzed as a separate exposure. Non-DNMT3A CHIP was defined from the maximum VAF across all available non-DNMT3A driver-gene columns. Participants with a non-DNMT3A clone could also harbour a DNMT3A clone because the exposure-specific models were not mutually exclusive. For continuous proteome-wide analyses, the maximum relevant VAF was transformed as LOG(1+100xVAF) and standardized within sex.

### Age-related variation in clone burden

The age dependence of mLOY, mLOX, non-DNMT3A CHIP and DNMT3A-mutant CHIP burden was modelled between ages 40 and 80 years. For each exposure, a linear model related clone burden to a natural cubic spline of age with four degrees of freedom and adjusted for smoking, recorded ethnicity, body-mass index (BMI), LDL cholesterol, hypertension, diabetes and the first ten genetic principal components. mLOY and mLOX percentage were analyzed on their original scale. Maximum non-DNMT3A and DNMT3A VAFs were analyzed after LOG(1+100xVAF) transformation and back-transformed for presentation. Curves are shown with 95% confidence intervals.

### Clinical outcomes and follow-up

Incident diagnoses were ascertained from UK Biobank longitudinal health-record data using International Classification of Diseases, 10th Revision (ICD-10) codes. The principal age-related disease composite comprised ischemic heart disease (I20–I25), heart failure (I50), stroke (I60–I64), Alzheimer’s disease (F00 or G30), pneumonia (J10–J18), chronic obstructive pulmonary disease (J43–J44), sepsis (A40–A41), chronic kidney disease (N18), type 2 diabetes (E11), osteoporosis (M80–M82) and polyarthritis (M05, M06 or M13). For multi-code outcomes, the earliest post-baseline incident event time was used. Participants without an event were censored at the longest available follow-up time among the component records. All-cause mortality was analyzed separately using the death indicator and corresponding follow-up time.

### Association of mosaic alterations with the age-related disease composite and mortality

Sex-stratified Cox proportional-hazards models estimated associations of dichotomous mLOY, mLOX, non-DNMT3A CHIP and DNMT3A-mutant CHIP with the age-related disease composite and all-cause mortality. Each exposure was fitted in a separate model. Unadjusted, age-adjusted and fully adjusted models were calculated. The principal estimates were from models adjusted for age at recruitment, smoking, recorded ethnicity, BMI, LDL cholesterol, hypertension, prevalent diabetes and first ten principal components of genetic ancestry. Models used Efron’s approximation for ties and complete observations for all included variables. Hazard ratios (HRs) are reported with 95% confidence intervals. Benjamini-Hochberg correction was applied within each outcome and model.

### Landmark analyses of temporal stability

To assess the temporal stability of associations with baseline clonal hematopoietic alterations, we repeated the fully adjusted sex-stratified Cox analyses in prespecified follow-up intervals of 0–3, 3–6 and 6–10 years. For intervals beginning after baseline, participants contributed risk time only if they remained under observation and were free of the respective disease-composite endpoint, or alive for the mortality analysis, at the start of the interval. Models used the same baseline exposure definitions, covariates, complete-case approach and Efron approximation for ties as the primary analysis. Benjamini–Hochberg correction was applied across the six sex–exposure tests within each outcome and follow-up interval. Temporal heterogeneity was assessed in a counting-process Cox model with interval-specific exposure coefficients, interval-stratified baseline hazards and participant-clustered robust covariance; equality of the three exposure coefficients was evaluated using a joint Wald test. Because clonal alterations were measured only at baseline, these analyses estimate the time-varying association of baseline clonal status and do not assess clone progression.

### Biomarker-derived measures of biological ageing

Associations with biological-age measures were assessed separately in men and women by ordinary least-squares regression. Each measure was modelled as a continuous outcome and each clonal alteration as a separate dichotomous exposure. Models were adjusted for age at recruitment, smoking, recorded ethnicity, BMI, LDL cholesterol, hypertension, diabetes and first ten principal components of genetic ancestry. Analyses required at least 250 complete observations and used two-sided tests with 95% confidence intervals.

### PhenoAge acceleration

PhenoAge was calculated as described previously using the published Levine coefficients^72^. The mortality-risk transformation was converted to PhenoAge using the published Gompertz parameters. PhenoAge acceleration was the residual from a sex-specific linear regression of PhenoAge on chronological age. Positive values indicated older biological age than expected.

### Klemera-Doubal biological age

Klemera-Doubal method (KDM) biological age was as described previously^73^. For each biomarker, its association with chronological age was estimated in the complete biomarker training sample, with at least 5,000 observations required. The fitted slopes, intercepts and residual variability were combined using the KDM equation. KDM age acceleration was the residual from a sex-specific regression of KDM biological age on chronological age.

### Telomere age gap

Relative leukocyte telomere length was taken from the log-transformed, technical-factor-adjusted T/S ratio and standardized. The telomere age gap was defined as the residual from a sex-specific regression of standardized telomere length on chronological age. In the source analysis, lower residuals indicate shorter telomeres than expected for age.

### Twenty-protein age estimate

A 20-protein age model was constructed from ACRV1, AGRP, CDCP1, COL6A3, CXCL17, EDA2R, ELN, ENG, FSHB, GDF15, GFAP, IGDCC4, KLK3, KLK7, LECT2, LTBP2, NEFL, PODXL2, PTPRR and SCARF2. The panel was selected from a previously published proteomic age clock^24^. The prediction model was re-estimated in the UK Biobank proteomic subset. Five-fold out-of-fold extreme gradient boosting was used (seed 20260627; squared-error objective; learning rate 0.03; maximum depth 3; minimum child weight 5; row and column subsampling 0.85; L2 penalty 1; L1 penalty 0; up to 2,000 boosting rounds; early stopping after 30 rounds). Protein values were standardized using parameters estimated within each training fold. Proteomic age gap (ProtAgeGap20) was predicted age minus chronological age.

For the cumulative-incidence analyses, a high proteomic age gap was defined as a value at or above the sex-specific median. Participants with any prevalent component of the 11-disease composite were excluded. Within the high-gap stratum, the reference group had neither sex-chromosome loss nor non-DNMT3A CHIP. Comparison groups additionally had mLOY or mLOX, or non-DNMT3A CHIP. Participants with both sex-chromosome loss and non-DNMT3A CHIP were excluded from these mutually exclusive comparisons. Cumulative incidence was estimated as one minus the Kaplan–Meier survival estimate over 10 years. Adjusted Cox models used the principal covariate set described above, and Benjamini-Hochberg correction was applied across the two comparisons within each sex.

### Phenome-wide association analysis

The incident-disease phenome-wide association study included non-congenital ICD-10 outcomes from 13 chapters: infectious (A00–B99), blood (D50–D89), endocrine (E00–E90), mental and behavioural (F00–F99), nervous system (G00–G99), eye (H00–H59), ear (H60– H95), circulatory (I00–I99), respiratory (J00–J99), digestive (K00–K93), musculoskeletal (M00–M99), skin (L00–L99) and genitourinary (N00–N99) diseases. Outcome-specific variables excluded prevalent diagnoses. Only outcomes with at least 200 incident events were analysed.

Within each sex, separate fully adjusted Cox models were fitted for mLOY or mLOX, non-DNMT3A CHIP and DNMT3A-mutant CHIP. Models used Efron’s method for ties and adjusted for age at recruitment, smoking, recorded ethnicity, BMI, LDL cholesterol, hypertension, diabetes and first ten principal components of genetic ancestry. Benjamini-Hochberg *q* values were calculated across all tested ICD outcomes separately within each sex and exposure. *q*<0.05 defined statistical significance. An outcome was labelled shared when it was significant for mLOY or mLOX and for either non-DNMT3A or DNMT3A-mutant CHIP.

### Model-based attributable disease burden

Exposure-specific model-based attributable fractions among exposed participants were estimated at a 10-year horizon separately for each sex, exposure and outcome. Fully adjusted Cox proportional-hazards models included the focal clonal alteration and age at recruitment, smoking, recorded ethnicity, body-mass index, LDL cholesterol, hypertension, prevalent diabetes and the first ten principal components of genetic ancestry. Each clonal alteration was evaluated in a separate exposure-specific model.

For each participant, the predicted 10-year risk under the fitted model was calculated using the estimated baseline cumulative hazard and the participant-specific linear predictor, with the focal exposure and all covariates retained at their observed values. For participants carrying the focal exposure, a corresponding predicted risk under an unexposed reference scenario was calculated by setting the focal exposure to zero while retaining all other covariates at their observed values.

For each exposed participant, the exposure-associated risk difference was calculated as the predicted risk under the observed exposure status minus the predicted risk under the unexposed reference scenario. These risk differences were summed across exposed participants and divided by the sum of their predicted risks under the observed exposure status. The resulting value was multiplied by 100 and reported as the attributable fraction among exposed participants.

AF therefore represents the proportion of predicted 10-year events among participants carrying the focal exposure that was associated with that exposure under the fitted model. Signed risk differences were retained without truncation. negative estimates indicate a lower predicted risk under the observed exposure model and were not interpreted as attributable harm.

### Plasma proteomic profiling and proteome-wide association analysis

The proteomic analyses used imputed, z-standardized UK Biobank Olink plasma-protein data generated through the UK Biobank Pharma Proteomics Project, which has been described previously^74^. A total of 46,324 participants had proteomic measurements available. Protein-wise analyses required at least 90% non-missing measurements, non-zero variance and at least 100 complete participant observations. 2,846 proteins were represented in the final proteome-wide result files. Olink plate was included as an additional technical covariate.

Proteome-wide associations were estimated separately in men and women using limma linear models with empirical-Bayes variance moderation. The primary continuous profiles were mLOY in men, mLOX in women, non-DNMT3A CHIP in men and women, and, for secondary analyses, DNMT3A-, TET2- and ASXL1-mutant CHIP. Each exposure was fitted separately and adjusted for age at recruitment, smoking, recorded ethnicity, BMI, LDL cholesterol, hypertension, diabetes, first ten principal components of genetic ancestry and Olink plate. Benjamini-Hochberg correction was applied across all proteins within each sex– exposure model. *q*<0.05 defined a significant protein association.

### Blood-cell-composition sensitivity analysis

The four primary proteome-wide association profiles were repeated with additional adjustment for neutrophil, lymphocyte, monocyte, eosinophil and basophil percentages and platelet count. These covariates were z-standardized for numerical stability. all other eligibility criteria, exposure transformations, covariates, Olink plate adjustment, limma empirical-Bayes inference and sex–exposure-specific Benjamini–Hochberg correction were unchanged. Because the five leukocyte percentages are compositional and sum to approximately 100%, a further sensitivity model omitted basophil percentage while retaining the same participants and all other covariates. Individual leukocyte-percentage coefficients were not interpreted.

### Proteomic landscapes

#### UMAP representation

Proteins associated at *q*<0.05 with at least one of the four primary profiles (men-mLOY, men- non-DNMT3A CHIP, women-mLOX and women-non-DNMT3A CHIP) were included in the proteomic landscape. For each protein, the dominant profile was the profile with the largest absolute signed −LOG_10_(*q*) value, and upregulated and downregulated proteins were embedded separately. The four-profile signed −LOG_10_(*q*) matrix was truncated to the interval −10 to 10, missing values were set to zero and dimensions were standardized. UMAP was run with 25 neighbours, minimum distance 0.20, cosine distance and seed 123. The UMAP was used as an exploratory visualization of association-profile similarity and not as inferential evidence.

#### Definition of UMAP domains

Regions were reconstructed separately in the upregulated and downregulated UMAPs using density-based spatial clustering (DBSCAN; ε=1.10; minimum points=8). Points labelled as noise were assigned to the nearest region centroid. The second upregulated region was subsequently split into mLOY-dominant and CHIP-dominant subregions: proteins whose dominant profile was mLOY or CHIP served as anchors, and remaining proteins were assigned to the nearest anchor centroid. The six domains used downstream were LOX- upregulated (14 proteins), mLOY-upregulated (132), CHIP-upregulated (89), CHIP-downregulated (35), mLOY-downregulated (47) and LOX-downregulated (8).

##### Clustering and functional annotation of protein profiles

A second, independent clustering analysis was performed in the original four-dimensional association-profile space rather than on UMAP coordinates. The input comprised the signed −LOG_10_(*q*) values for the four primary sex-exposure profiles. Proteins significant in at least one profile were eligible. Missing values were set to zero. The 325 retained proteins were hierarchically clustered using correlation distance and average linkage. Cluster solutions from k=4 to k=12 were evaluated by silhouette width. The prespecified k=8 solution was post-processed by merging clusters containing fewer than five proteins with the nearest cluster, yielding six clusters (50, 133, 84, 33, 12 and 13 proteins).

Functional labels were assigned using a pre-defined analysis plan. Overrepresentation analyses used Gene Ontology Biological Process and Reactome, with all proteins measured in the corresponding Olink analysis as the primary background. A competitive analysis restricted the background to proteins admitted to profile clustering to assess whether a term distinguished one cluster from the other association-profile clusters. Redundant Gene Ontology terms were collapsed using semantic similarity, and overlapping Reactome terms were reduced on the basis of shared cluster genes. Functional enrichment was independently repeated through the STRING application programming interface using the same assay-specific background. Robustness was assessed by comparing the original and blood-cell-adjusted cluster solutions, calculating silhouette widths, testing 10,000 randomly sampled protein sets matched to each cluster size, and repeating enrichment after removing each protein in turn from the two smallest clusters. Cluster names were retained only when the biological theme was supported across databases or sensitivity analyses; otherwise, broader descriptive labels were used.

This procedure yielded the labels ‘haematopoietic growth-factor signalling’, ‘neutrophil degranulation and extracellular-matrix remodelling’, ‘myeloid immune regulation’, ‘chemotactic, haemostatic and extracellular-matrix signalling’, ‘cytotoxic lymphocyte/NK-cell response’ and ‘B-cell/cytokine-associated signalling’. These names are functional annotations of association-profile clusters and do not imply that every member participates in the named process or that the cluster represents a causal pathway. The B-cell/cytokine-associated label was treated as exploratory. The heatmap displayed up to ten proteins per cluster with q<0.05 in at least one profile, ranked by statistical evidence, and sex-specific mirror plots displayed up to 15 proteins per cluster.

### External genetic validation of protein–disease associations

Causal relationships between domain proteins and disease outcomes were investigated with two-sample Mendelian-randomization (MR). Protein cis-quantitative trait loci (cis-pQTLs) were derived from UK Biobank Pharma Proteomics Project summary statistics of circulating protein levels measured with the Olink platform^74^. For 273 out of 325 domain proteins (mLOY-upregulated N=103, CHIP-upregulated N=76, mLOX-upregulated N=13, mLOY-downregulated N=42, CHIP-downregulated N=31 and mLOX-downregulated N=8) at least one cis-pQTL (range 1-40) meeting defined criteria (*P*<5E-6, +/-50 kilobases up-/downstream of gene borders) was identified. The relaxed *P*-value threshold compared to the genome-wide threshold of P<5E-8 in combination with the narrow window around gene borders allowed to optimize power while ensuring inclusion of biologically highly relevant genetic instruments and reducing the risk of horizontal pleiotropy. Cis-pQTLs were used as instrumental variables (IVs) in the MR analysis. If more than one cis-pQTL was identified for a given gene/protein, variants were clumped to retain only variants in low linkage disequilibrium (LD; r²<0.1), i.e., largely independent variants, in the analysis. LD structure information was included in all multi-IV analyses to account for variant correlation by including linkage information from the European panel of the 1000 Genomes project (phase 3). All IVs included had an F-statistic > 10, indicating strong instruments.

Disease outcomes included coronary disease, heart failure, stroke, pneumonia, chronic obstructive pulmonary disease (COPD), sepsis, type 2 diabetes, chronic kidney disease, osteoporosis, polyarthritis and Alzheimer disease. Corresponding summary statistics for each outcome were obtained from the FinnGen cohort^75^ (release 12) of over 500,000 individuals. For COPD and polyarthritis, two corresponding summary statistics were available, i.e., COPD_early and COPD_later, as well as monoarthritis and polyarthritis, respectively. Thus, both were included in the analysis and *P*-values were pooled post-hoc for downstream analysis and visualization using the aggregated Cauchy association test (ACAT) implemented in the ACAT package^76^.

All genetic data were derived from individuals of European ancestry, and there was no overlap between the exposure and outcome study cohorts. Data was harmonized with the harmonise_data function of TwoSampleMR^77^. Multi-allelic and palindromic variants were excluded.

Single-instrument analyses were performed with the Wald-ratio test. Analyses with 2-3 IVs for a given protein were conducted with the fixed-effects inverse-variance-weighted (IVW) method. If more than three IVs were present, we used a random-effects IVW model.

Sensitivity analyses with MR-Egger were performed for all proteins with >3 IVs and nominal significance (*P*<0.05) in the primary analysis. Genetics associations with >3 IVs were considered robust if the effect estimates were directionally concordant and significant in primary and sensitivity analyses and did not have a significant heterogeneity statistic. To obtain a global estimate of genetic association for each protein domain-outcome pair, *P* values were combined post-hoc within each domain using ACAT^76^. MR analyses were performed using the TwoSampleMR^77^ and MendelianRandomization^78^ packages in R. For reproducibility of results, MR analyses were implemented as a snakemake^79^ pipeline and run in a customized docker container.

## Statistics and reproducibility

All tests were two-sided. Effect estimates are reported with 95% confidence intervals. Continuous clonal-burden and proteomic-domain variables were standardized where stated, so their coefficients represent a one-standard-deviation difference. Unless otherwise specified, the common adjustment set comprised age at recruitment, smoking, recorded ethnicity, BMI, LDL cholesterol, hypertension, diabetes and first ten principal components of genetic ancestry. Overall proteome-wide models additionally included Olink plate. False-discovery-rate control used the Benjamini–Hochberg procedure within the families specified for each analysis. The nominal significance threshold was *P*<0.05 and the multiple-testing threshold was *q*<0.05.

Analyses were performed in R using, among other packages, survival, limma, broom, splines, uwot, dbscan, ComplexHeatmap, ggplot2, dplyr, tidyr, readr, data.table, cluster, clusterProfiler, ReactomePA, GOSemSim, httr and jsonlite, and xgboost. The analysis scripts contain fixed seeds for the UMAP (123), protein-profile clustering fallbacks (123) and proteomic-age model (20260627).

## Data availability

UK Biobank data are available to bona fide researchers through the UK Biobank access procedure. These analyses were conducted under application 170256.

## References

1. Chang, A.Y., Skirbekk, V.F., Tyrovolas, S., Kassebaum, N.J. & Dieleman, J.L. Measuring population ageing: an analysis of the Global Burden of Disease Study 2017. Lancet Public Health 4, e159–e167 (2019).

2. Lopez-Otin, C., Blasco, M.A., Partridge, L., Serrano, M. & Kroemer, G. The hallmarks of aging. Cell 153, 1194–1217 (2013).

3. Evans, M.A. & Walsh, K. Clonal hematopoiesis, somatic mosaicism, and age- associated disease. Physiol Rev 103, 649–716 (2023).

4. Forsberg, L.A., Gisselsson, D. & Dumanski, J.P. Mosaicism in health and disease - clones picking up speed. Nat Rev Genet 18, 128–142 (2017).

5. Yousefzadeh, M.J., et al. An aged immune system drives senescence and ageing of solid organs. Nature 594, 100–105 (2021).

6. Steensma, D.P., et al. Clonal hematopoiesis of indeterminate potential and its distinction from myelodysplastic syndromes. Blood 126, 9–16 (2015).

7. Machiela, M.J., et al. Female chromosome X mosaicism is age-related and preferentially affects the inactivated X chromosome. Nat Commun 7, 11843 (2016).

8. Forsberg, L.A., et al. Mosaic loss of chromosome Y in peripheral blood is associated with shorter survival and higher risk of cancer. Nat Genet 46, 624–628 (2014).

9. Thompson, D.J., et al. Genetic predisposition to mosaic Y chromosome loss in blood. Nature 575, 652–657 (2019).

10. Jaiswal, S., et al. Age-related clonal hematopoiesis associated with adverse outcomes. N Engl J Med 371, 2488–2498 (2014).

11. Genovese, G., et al. Clonal hematopoiesis and blood-cancer risk inferred from blood DNA sequence. N Engl J Med 371, 2477–2487 (2014).

12. Jaiswal, S., et al. Clonal Hematopoiesis and Risk of Atherosclerotic Cardiovascular Disease. N Engl J Med 377, 111–121 (2017).

13. Jaiswal, S. & Ebert, B.L. Clonal hematopoiesis in human aging and disease. Science 366(2019).

14. Jaiswal, S. Clonal hematopoiesis and nonhematologic disorders. Blood 136, 1606–1614 (2020).

15. Forsberg, L.A. Loss of chromosome Y (LOY) in blood cells is associated with increased risk for disease and mortality in aging men. Hum Genet 136, 657–663 (2017).

16. Sano, S., et al. Hematopoietic loss of Y chromosome leads to cardiac fibrosis and heart failure mortality. Science 377, 292–297 (2022).

17. Saw, W.Y., et al. Mosaic loss of Y chromosome associates with lung function, emphysema, and epigenetic aging. Am J Respir Crit Care Med 212, 1483–1494 (2026).

18. Liu, A., et al. Genetic drivers and cellular selection of female mosaic X chromosome loss. Nature 631, 134–141 (2024).

19. Robertson, N.A., et al. Age-related clonal haemopoiesis is associated with increased epigenetic age. Curr Biol 29, R786–R787 (2019).

20. Nachun, D., et al. Clonal hematopoiesis associated with epigenetic aging and clinical outcomes. Aging Cell 20, e13366 (2021).

21. Yu, Z., et al. Human plasma proteomic profile of clonal hematopoiesis. Nat Commun 16, 11688 (2025).

22. Weyrich, M., et al. Loss of Y chromosome: proteomic signatures in human cardiovascular disease. Eur Heart J 46, 5237–5239 (2025).

23. Hubbard, A.K., et al. Serum biomarkers are altered in UK Biobank participants with mosaic chromosomal alterations. Hum Mol Genet 32, 3146–3152 (2023).

24. Argentieri, M.A., et al. Proteomic aging clock predicts mortality and risk of common age-related diseases in diverse populations. Nat Med 30, 2450–2460 (2024).

25. Kim, P.G., et al. Dnmt3a-mutated clonal hematopoiesis promotes osteoporosis. J Exp Med 218(2021).

26. Kessler, M.D., et al. Common and rare variant associations with clonal haematopoiesis phenotypes. Nature 612, 301–309 (2022).

27. Wang, H., et al. Clonal hematopoiesis driven by mutated DNMT3A promotes inflammatory bone loss. Cell 187, 3690–3711 e3619 (2024).

28. Zhao, K., et al. Association of Clonal Hematopoiesis With Incident, Late-Onset, Seropositive Rheumatoid Arthritis. Arthritis Rheumatol (2026).

29. Koh, Y., Tengesdal, I.W. & Jaiswal, S. Clonal Hematopoiesis in Nonmalignant Disease: Functional Consequences of Mutated Immune Cells by Clonal Hematopoiesis in the Diseased Tissue. Annu Rev Pathol 21, 19–36 (2026).

30. Moaddel, R., et al. Proteomics in aging research: A roadmap to clinical, translational research. Aging Cell 20, e13325 (2021).

31. Kuo, C.L., et al. Proteomic aging clock (PAC) predicts age-related outcomes in middle-aged and older adults. Aging Cell 23, e14195 (2024).

32. Kuo, C.L., et al. A proteomic signature of healthspan. Proc Natl Acad Sci U S A 122, e2414086122 (2025).

33. Robinson, O., et al. Associations of proteomic age clocks with lifestyle risk factors, incident chronic diseases and mortality in two European cohorts. Nat Aging 6, 1437-1451 (2026).

34. Solleti, S.K., et al. Airway epithelial cell PPARgamma modulates cigarette smoke- induced chemokine expression and emphysema susceptibility in mice. Am J Physiol Lung Cell Mol Physiol 309, L293–304 (2015).

35. Almansa, R., et al. Critical COPD respiratory illness is linked to increased transcriptomic activity of neutrophil proteases genes. BMC Res Notes 5, 401 (2012).

36. Gong, L. & Si, M.S. SLIT3-mediated fibroblast signaling: a promising target for antifibrotic therapies. Am J Physiol Heart Circ Physiol 325, H1400–H1411 (2023).

37. Giannandrea, M. & Parks, W.C. Diverse functions of matrix metalloproteinases during fibrosis. Dis Model Mech 7, 193–203 (2014).

38. Sakamoto, N., et al. HSP47: A Therapeutic Target in Pulmonary Fibrosis. Biomedicines 11(2023).

39. Razzaque, M.S., Le, V.T. & Taguchi, T. Heat shock protein 47 and renal fibrogenesis. Contrib Nephrol 148, 57–69 (2005).

40. Wang, Y., et al. The role of matrix metalloproteinase 9 in fibrosis diseases and its molecular mechanisms. Biomed Pharmacother 171, 116116 (2024).

41. Chen, Y., et al. Mitochondrial Metabolic Reprogramming by CD36 Signaling Drives Macrophage Inflammatory Responses. Circ Res 125, 1087–1102 (2019).

42. Chen, Y., Zhang, J., Cui, W. & Silverstein, R.L. CD36, a signaling receptor and fatty acid transporter that regulates immune cell metabolism and fate. J Exp Med 219(2022).

43. Kavurma, M.M. & Khachigian, L.M. Signaling and transcriptional control of Fas ligand gene expression. Cell Death Differ 10, 36–44 (2003).

44. Salminen, A., Kaarniranta, K. & Kauppinen, A. Regulation of longevity by FGF21: Interaction between energy metabolism and stress responses. Ageing Res Rev 37, 79–93 (2017).

45. Gliniak, C.M., et al. FGF21 promotes longevity in diet-induced obesity through metabolic benefits independent of growth suppression. Cell Metab 37, 1547–1567 e1546 (2025).

46. Weyrich, M., et al. Independent and Dose-Dependent Contributions of Clonal Hematopoiesis and Mosaic Loss of Y to Incident Heart Failure. Eur J Heart Fail (2026).

47. Weyrich, M., et al. Loss of Y Chromosome and Cardiovascular Events in Chronic Kidney Disease. Circulation 150, 746–757 (2024).

48. Arai, Y., et al. Hematopoietic loss of Y chromosome activates immune checkpoints and contributes to impaired senescent cell clearance and renal disease. Sci Transl Med 17, eadv4071 (2025).

49. Bjurling, J., et al. Mosaic loss of chromosome Y in blood is associated with male susceptibility for idiopathic pulmonary fibrosis. Commun Med (Lond) 5, 246 (2025).

50. Valasarajan, C., et al. Loss of Y Chromosome Associates With Lung and Cardiac Dysfunction in COPD. Compr Physiol 16, e70142 (2026).

51. Leal Rojas, I.M., et al. Human Blood CD1c(+) Dendritic Cells Promote Th1 and Th17 Effector Function in Memory CD4(+) T Cells. Front Immunol 8, 971 (2017).

52. Fang, X.Y., Xu, W.D., Pan, H.F., Leng, R.X. & Ye, D.Q. Novel insights into Tim-4 function in autoimmune diseases. Autoimmunity 48, 189–195 (2015).

53. Dietrich, J., Cella, M., Seiffert, M., Buhring, H.J. & Colonna, M. Cutting edge: signal- regulatory protein beta 1 is a DAP12-associated activating receptor expressed in myeloid cells. J Immunol 164, 9–12 (2000).

54. Takahashi, M., et al. Human CD300C delivers an Fc receptor-gamma-dependent activating signal in mast cells and monocytes and differs from CD300A in ligand recognition. J Biol Chem 288, 7662–7675 (2013).

55. Zhang, J., et al. Leukocyte immunoglobulin-like receptors in human diseases: an overview of their distribution, function, and potential application for immunotherapies. J Leukoc Biol 102, 351–360 (2017).

56. Wang, T., Tian, J. & Jin, Y. VCAM1 expression in the myocardium is associated with the risk of heart failure and immune cell infiltration in myocardium. Sci Rep 11, 19488 (2021).

57. Nemeth, K., et al. The role of osteoclast-associated receptor in osteoimmunology. J Immunol 186, 13–18 (2011).

58. Etzerodt, A. & Moestrup, S.K. CD163 and inflammation: biological, diagnostic, and therapeutic aspects. Antioxid Redox Signal 18, 2352–2363 (2013).

59. Ivetic, A., Hoskins Green, H.L. & Hart, S.J. L-selectin: A Major Regulator of Leukocyte Adhesion, Migration and Signaling. Front Immunol 10, 1068 (2019).

60. Hajishengallis, G. & Chavakis, T. Inflammageing and clonal haematopoiesis interplay and their impact on human disease. Nat Rev Mol Cell Biol 27, 377–393 (2026).

61. Pastoret, C., et al. Diagnostic criteria for NK cell large granular lymphocyte leukemia: validation through a multicentric international study. Blood Adv 10, 642–653 (2026).

62. Schreeder, D.M., Pan, J., Li, F.J., Vivier, E. & Davis, R.S. FCRL6 distinguishes mature cytotoxic lymphocytes and is upregulated in patients with B-cell chronic lymphocytic leukemia. Eur J Immunol 38, 3159–3166 (2008).

63. Bund, D., Mayr, C., Kofler, D.M., Hallek, M. & Wendtner, C.M. Human Ly9 (CD229) as novel tumor-associated antigen (TAA) in chronic lymphocytic leukemia (B-CLL) recognized by autologous CD8+ T cells. Exp Hematol 34, 860–869 (2006).

64. Daga, S., et al. High GPR56 surface expression correlates with a leukemic stem cell gene signature in CD34-positive AML. Cancer Med 8, 1771–1778 (2019).

65. Aubert, A., Jung, K., Hiroyasu, S., Pardo, J. & Granville, D.J. Granzyme serine proteases in inflammation and rheumatic diseases. Nat Rev Rheumatol 20, 361–376 (2024).

66. Takeuchi, A., et al. CRTAM determines the CD4+ cytotoxic T lymphocyte lineage. J Exp Med 213, 123–138 (2016).

67. Kline, D.E., et al. CD8alpha(+) Dendritic Cells Dictate Leukemia-Specific CD8(+) T Cell Fates. J Immunol 201, 3759–3769 (2018).

68. Kucuksezer, U.C., et al. The Role of Natural Killer Cells in Autoimmune Diseases. Front Immunol 12, 622306 (2021).

69. Sudlow, C., et al. UK biobank: an open access resource for identifying the causes of a wide range of complex diseases of middle and old age. PLoS Med 12, e1001779 (2015).

70. Loh, P.R., et al. Insights into clonal haematopoiesis from 8,342 mosaic chromosomal alterations. Nature 559, 350–355 (2018).

71. Jakubek, Y.A., et al. Genomic and phenotypic correlates of mosaic loss of chromosome Y in blood. Am J Hum Genet 112, 276–290 (2025).

72. Levine, M.E., et al. An epigenetic biomarker of aging for lifespan and healthspan. Aging (Albany NY) 10, 573–591 (2018).

73. Klemera, P. & Doubal, S. A new approach to the concept and computation of biological age. Mech Ageing Dev 127, 240–248 (2006).

74. Sun, B.B., et al. Plasma proteomic associations with genetics and health in the UK Biobank. Nature 622, 329–338 (2023).

75. Kurki, M.I., et al. FinnGen provides genetic insights from a well-phenotyped isolated population. Nature 613, 508–518 (2023).

76. Liu, Y., et al. ACAT: A Fast and Powerful p Value Combination Method for Rare- Variant Analysis in Sequencing Studies. Am J Hum Genet 104, 410–421 (2019).

77. Hemani, G., et al. The MR-Base platform supports systematic causal inference across the human phenome. Elife 7(2018).

78. Yavorska, O.O. & Burgess, S. MendelianRandomization: an R package for performing Mendelian randomization analyses using summarized data. Int J Epidemiol 46, 1734–1739 (2017).

79. Koster, J. & Rahmann, S. Snakemake-a scalable bioinformatics workflow engine. Bioinformatics 34, 3600 (2018).

